# Effect of levodopa treatment on gait in older adults with mild parkinsonian signs

**DOI:** 10.64898/2026.06.04.26354926

**Authors:** Chatkaew Pongmala, Stiven Roytman, Miriam van Emde Boas, Robert Vangel, Caterina Rosano, Nicolaas Bohnen

**Affiliations:** Department of Radiology, University of Michigan, Ann Arbor, MI 48105-9755, USA; Morris K. Udall Center of Excellence for Parkinson’s Disease Research, University of Michigan, Ann Arbor, MI, USA; Department of Epidemiology, University of Pittsburgh, Pittsburgh, PA, USA; Neurology Service and GRECC, VA Ann Arbor Healthcare System, Ann Arbor, MI, USA

## Abstract

**Background:** Slow walking in older adults with mild parkinsonian signs (MPS) is a complex, multifactorial phenomenon arising from the cumulative burden of subclinical age-associated pathologies. This decline reflects age-associated neuronal loss in the dopaminergic system. A recent study suggests that levodopa treatment may enhance gait parameters. The goal of this small pilot study is to explore the effect of levodopa treatment on slow walking gait in older adults with MPS.

**Method:** This study was a randomized, placebo-controlled clinical pilot trial. Slow walking older adults without clinical evidence of PD were recruited and randomized into 2 groups (active treatment group or placebo control group). Participants in the active group were pre-treated with carbidopa for three days, followed by carbidopa-levodopa for seven days. Spatiotemporal gait parameters were evaluated at baseline and post-intervention.

**Results:** Gait factor analysis identified three main factors explaining gait characteristics at baseline, which included “gait efficiency”, “gait rhythmicity”, and “gait turning”.No effect of treatment was observed in the placebo group (β=0.111, p=0.616), no group difference was observed between the placebo and active group at baseline (β=0.310, p=0.547), but a strong trend for a treatment-related increase was observed in the active treatment group (β=0.506, p=0.076).

**Conclusion:** Our preliminary data suggest that sustained levodopa treatment (one week) in conjunction with carbidopa pre-treatment and concomitant carbidopa supplementation is feasible in slow walking older adults with MPS. Moreover, the data indicate potential efficacy, showing improvements in cadence, and step durations.

## Background

Mild parkinsonian signs (MPS) are common in aging, with an estimated prevalence up to 50%^1^. MPS have been described during the neurological examination of older adults in the absence of specific parkinsonian syndromes^1^. The typical clinical manifestations of MPS include bradykinesia, rigidity, and/or postural instability and gait disturbance (PIGD) motor features^1–4^. MPS are often associated with risks of fear of falling, Parkinson’s disease, dementia with Lewy bodies (DLB), or cognitive decline or may be seen in the setting of depression with psychomotor slowing^4–6^. The majority of older adults with MPS demonstrate signs of PIGD and bradykinesia, while few have rest tremor^3,4,7,8^.

Previous studies showed that a potential contributor to MPS in older adults is a decline in age-associated dopamine with lower levels of dopamine nerve terminals, or dopamine rD1/D2 receptor densities^9–12^. There is also increasing evidence that Levodopa improves gait in older adults with MPS^13–16^. A recent open-label study from our center showed that levodopa treatment improved cadence, pace, and time spent in the double support phase in older adults with MPS^17^. However, open label studies may suffer from placebo effects. Presently, there is a lack of randomized controlled clinical trials on the effects of Levodopa on slow walking gait in older adults with MPS. The goal of this small placebo-controlled phase 1b pilot study is to explore the effect of levodopa treatment on slow walking gait in older adults with MPS. We hypothesized that levodopa treatment can improve slow walking gait in older adults with MPS, in particular cadence, pace and time spent in double support.

## Method

This randomized controlled clinical phase 1b pilot study included older adults with MPS recruited via the UMHealthResearch website between March and May 2024. The study was part of a NIH-funded study on neurobiological drivers of mobility resilience (clinicaltrials.gov NCT06219915 and NCT06587217). The study was approved by the medical ethics committee at the University of Michigan (HUM00156490) and informed consent was obtained from all study participants. This study was performed in line with the principles of the Declaration of Helsinki.

Older adults, aged 60 years and over, who affirmatively answered whether they have noticed slowing of gait over at least a year, and who did not have a diagnosis of PD or other neurological diseases, were recruited and screened for slow walking speed (< 1 m/s) and/or MPS features in clinical examination (predominantly features on postural instability or gait impairment). Patients with clear clinical evidence of Parkinson’s disease were excluded from the study.

70 individuals were contacted for the study. Of these, 16 completed pre-screening and were enrolled in the study. Two participants screen-failed due to an absence of MPS (walking speed > 1 m/s). In total, 14 participants were randomized and completed the study intervention. See **Table 1** for baseline descriptive statistics.

**Table 1.** Descriptive statistics by intervention group at baseline.

|  | Placebo (n=5) |  | L-dopa (n=9) |  | P-value |  |
| --- | --- | --- | --- | --- | --- | --- |
|  | Mean | SD | Mean | SD | 1-tailed | 2-tailed |
| Age | 77.2 | 6.3 | 77.4 | 5.8 | 0.47 | 0.94 |
| Sex (M/F) | 2/3 |  | 6/3 |  | 0.34 | 0.58 |
| MDS-UPDRS III | 30.3 | 10.73 | 21.39 | 5.24 | 0.03 | 0.06 |
| Velocity (m/s) | 0.89 | 0.12 | 0.89 | 0.08 | 0.49 | 0.99 |
| Cadence (steps/min) | 104.29 | 10.68 | 104.94 | 7.87 | 0.45 | 0.90 |
| Stride length (m) | 1.02 | 0.14 | 1.01 | 0.10 | 0.45 | 0.90 |
| Double support (%GCT) | 25.93 | 1.75 | 25.48 | 2.74 | 0.37 | 0.75 |
| Gait cycle duration (s) | 1.16 | 0.11 | 1.15 | 0.08 | 0.42 | 0.83 |
| Single limb support (%GCT) | 37.07 | 0.81 | 37.22 | 1.35 | 0.41 | 0.82 |
| Stance (%GCT) | 62.99 | 0.97 | 62.69 | 1.40 | 0.34 | 0.69 |
| Step duration (s) | 0.58 | 0.05 | 0.58 | 0.04 | 0.41 | 0.82 |
| Swing (%GCT) | 37.01 | 0.97 | 37.31 | 1.40 | 0.34 | 0.69 |
| Arm swing velocity (degrees/s) | 116.15 | 24.67 | 153.91 | 39.60 | 0.04 | 0.08 |
| Arm ROM (degrees) | 23.06 | 4.54 | 32.36 | 10.07 | 0.04 | 0.08 |
| Turn angle (degrees) | 175.46 | 6.18 | 172.57 | 36.05 | 0.43 | 0.86 |
| Turn duration (s) | 2.72 | 0.41 | 2.66 | 0.62 | 0.43 | 0.86 |
| Turn Velocity (degrees/s) | 120.02 | 17.65 | 131.04 | 25.50 | 0.21 | 0.41 |

This study was a randomized placebo-controlled clinical pilot trial. Prior to any assessments, the participants will be randomized into 2 groups. The groups are the experimental or active group and the control or placebo groups.The experimental or active group received a carbidopa-levodopa treatment while the control group received a placebo treatment.

All participants were assessed for parkinsonian signs on day 0. Clinical assessments were completed for qualified participants. To reduce the risk of nausea and vomiting, participants in the active group were pre-treated with 25 mg of carbidopa monotherapy orally (PO) three times a day (TID) for three days prior to starting the carbidopa-levodopa treatment portion of the study (days 1-3). All participants in the active group continued to take 25 mg carbidopa PO TID for the entire treatment duration. A low dose of carbidopa-levodopa immediate release (25/100 mg 1 tablet PO TID) was used for the first three days of treatment (days 4-6). If no side-effects occurred, the dose was then increased to 1.5 tablets PO TID for days 7-10 (figure 1). All medications were prepared and dispensed by the Research Pharmacy at the University of Michigan. At the mean time, all participants in the control group will follow the same protocol, however, the medications they received will be a placebo.

**Figure 1.**
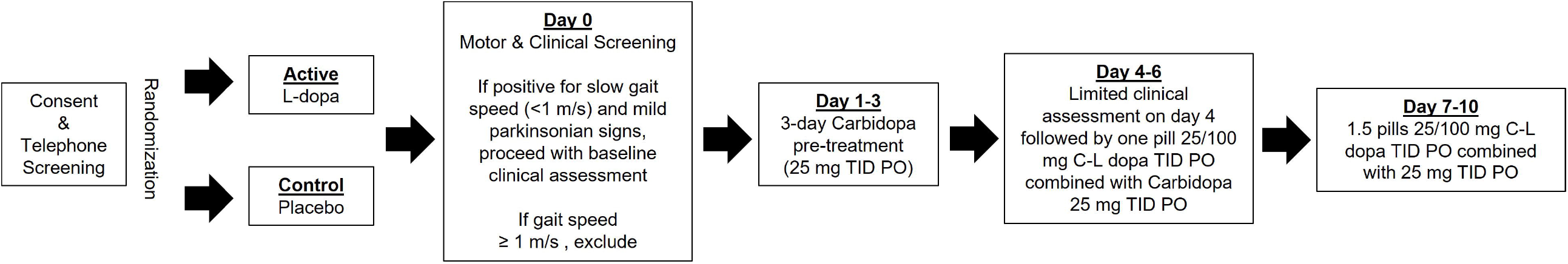
study timeline.

Demographic information was obtained for all participants including age, gender, and years of education. Motor symptom evaluation was performed using the MDS-UPDRS Part III by a movement disorder expert. Eligible participants completed the Instrumented Stand and Walk test (iSAW) APDM Mobility Lab^™^ protocol with six inertial measurement units (IMU) attached at the sternum, lumbar, feet, and wrists. Motor testing was completed at baseline, post-carbidopa pre-treatment, and post carbidopa-levodopa treatment visits.

For the iSAW protocol, participants stand still for thirty seconds, walk 7 m (indicated by markings on the floor), make a 180° turn around a cone, then walk 7 m back to the initial standing point.

**Figure 2.**
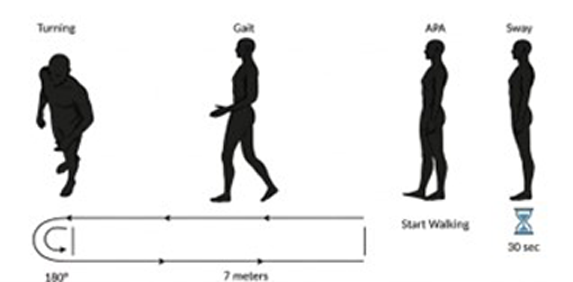
ISAW protocol.

### Statistical analysis

We compared groups at baseline on demographic and gait parameters using the Chi-square test for categorical data and independent samples-test for continuous variables. Factor analysis was performed on 14 gait variables (listed in table 1), to reduce feature dimensionality (see supplementary materials for description of factor analysis methods). A linear mixed model was used to predict each of the gait factor scores from an interaction between group and visit. Likelihood ratio test model comparisons were performed against a null model without an interaction effect, and coefficients of statistically significant or strongly trending models were reported. All tests were two-tailed with a P-value < 0.05. Statistical analyses were performed using R statistical software.

## Results

9 (3F/5M) were recruited in an experimental L-dopa treatment group (mean age 77.4 ± 5.8 yrs, and mean MDS-UPDRS III 21.39 ± 5.24), and 5 (3F/2M) were recruited in a control placebo group (mean age 77.2 ± 6.3 yrs, and mean MDS-UPDRS III 30.3 ± 10.73). Full descriptive statistics are presented in table 1.The study intervention was well tolerated. Mild side-effects included headache (2 affected), blood pressure fluctuations (1 affected), dizziness (2 affected), nausea (1 affected) and drowsiness (5 affected). There were no related moderate or serious adverse events.

### Gait Factor Analysis

Gait factor analysis identified a three main factors explaining gait characteristics at baseline, which included “gait efficiency” (variance explained = 38%), “gait rhythmicity” (variance explained = 29%), and “gait turning” (variance explained = 15%). Gait efficiency had strong positive loadings for swing/single-support time, and negative loadings for stance/double-support time, with higher factor scores considered more clinically favorable. Gait rhythmicity had strong positive loadings on cadence and arm range of motion and strong negative loadings on step/gait cycle duration, with higher factors scores considered more clinically favorable. Gait turning had only strong positive loadings on turn duration and angle, with lower factor scores considered more clinically favorable.

### Linear mixed model analysis

Full set of model coefficients for “gait rhythmicity” is presented in Table 2. No effect of treatment was observed in the placebo group (β=0.111, p=0.616), no group difference was observed between placebo and active group at baseline (β=0.310, p=0.547), but a strong trend for a treatment-related increase was observed in the active treatment group (β=0.506, p=0.076). Individual participant changes in gait rhythmicity are plotted by group in Figure 3, and demonstrate a consistent increase in all patients within the active treatment group, with heterogeneous changes observed in the placebo group. Given the potentially confounding effect of baseline group differences on motor symptom severity (MDS-UPDRS part III total score), the gait rhythmicity model was refitted with baseline visit motor symptom severity as an additional covariate. Presence of baseline motor symptom severity as a covariate did not substantially affect the coefficient estimate for the effect of treatment on the active group (β=0.506, p=0.076).

**Table 2.** Regression coefficients from the “gait rhythmicity” linear mixed model.

| Gait Rhythmicity |  |  |  |
| --- | --- | --- | --- |
| Predictors | Estimates | CI | p |
| (Intercept) | -0.418 | -1.262 – 0.427 | 0.316 |
| Visit [post] | 0.111 | -0.341 – 0.563 | 0.616 |
| Group [active] | 0.310 | -0.743 – 1.364 | 0.547 |
| Visit [post] ×<br>Group [active] | 0.506 | -0.057 – 1.069 | **0.076** |
| Random Effects |  |  |  |
| $\sigma^2$ | | 0.12 | |
| $\tau_{00}$ Subject | | 0.71 | |
| ICC |  | 0.86 |  |
| N<sub>Subject</sub> |  | 14 |  |
| Observations |  | 28 |  |
| Marginal R<sup>2</sup> / Conditional<br>R<sup>2</sup> |  | 0.145 / 0.878 |  |

*-Please insert **Figure 3** Individual participant changes in gait rhythmicity from pre-treatment to post-treatment by intervention group.here –*

**Figure 3.**
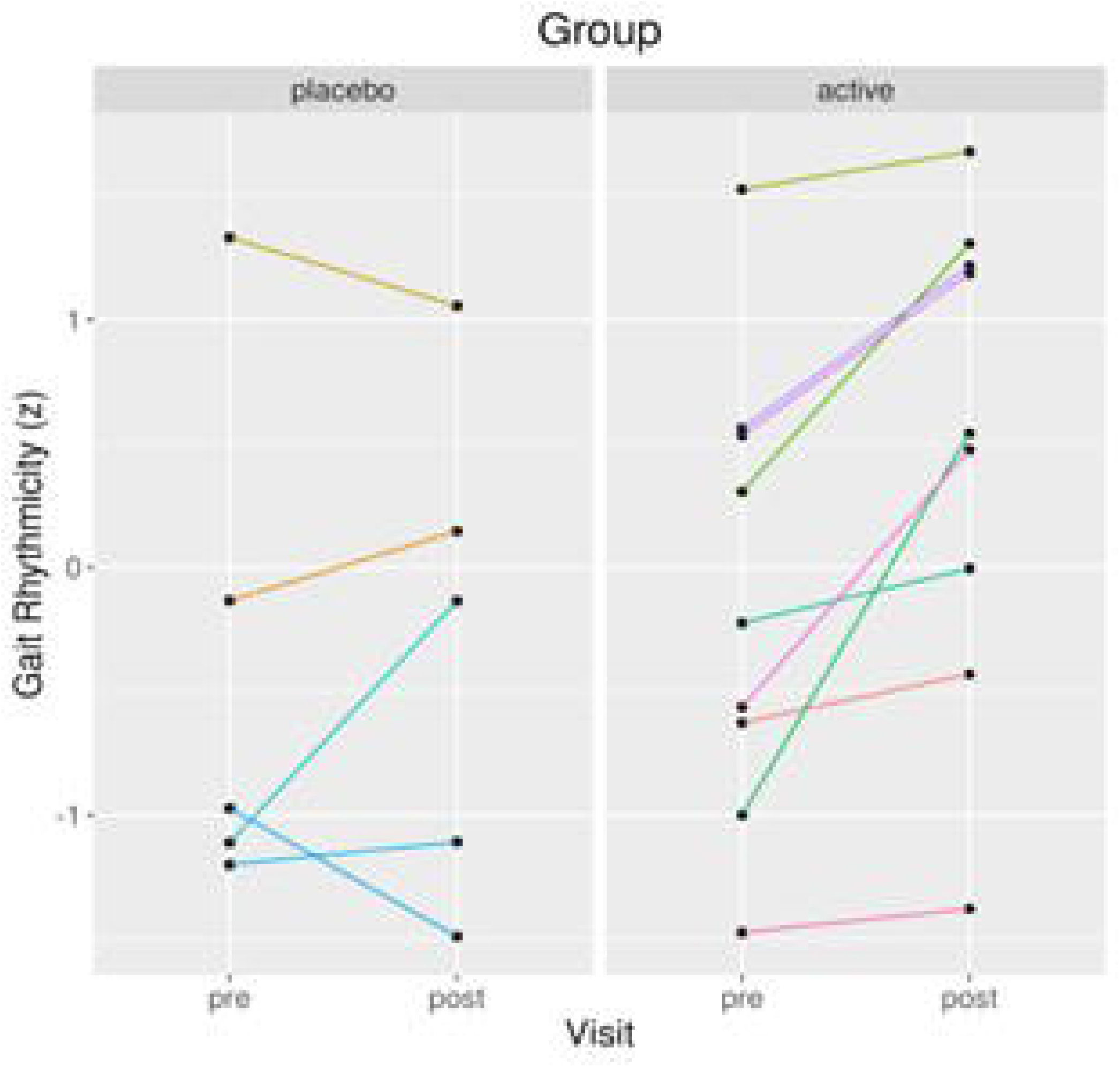
Individual participant changes in gait rhythmicity from pre-treatment to post-treatment by intervention group.

Given that gait rhythmicity failed to reach two-tailed statistical significance, we performed a power analysis with the *R simr* software library to determine a sample size which would be sufficient to detect significant effects of the magnitude observed in this study. At a total sample size of N=35 (placebo=13, active=22), the power to observe a significant group x visit interaction for gait rhythmicity was 82.5% (CI_95_= [80.0, 84.81]%).

## Discussion

Our study is a randomized placebo-controlled clinical phase 1b pilot trial investigating one week of sustained levodopa treatment in older adults with slow walking, aiming to examine the effect of levodopa treatment on slow walking gait in the elderly population. From a clinical perspective, our preliminary data suggest that sustained levodopa treatment (one week) in conjunction with carbidopa pre-treatment and concomitant carbidopa supplementation is feasible in slow walking older adults with MPS. Moreover,the data indicate potential efficacy, showing improvements in cadence, and step durations. Our findings are consistent with previous research on older adults with MPS^17^. These results provide valuable insights into the potential of using levodopa treatment to treat older adults with MPS in the future.

Slow walking in older adults with MPS is a complex, multifactorial phenomenon arising from the cumulative burden of subclinical age-associated pathologies. This decline typically reflects a confluence of nigrostriatal dopaminergic attrition, degenerative joint disease, sarcopenia, and cerebral white matter changes, which collectively undermine motor automaticity^18^. Recent evidence suggests that levodopa (L-DOPA) therapy may enhance gait parameters in this population by providing a “dopaminergic resilience” function, effectively buffering against these diverse physiological stressors^17^. Consequently, exogenous dopamine may serve as a critical substrate for maintaining mobility and potentially reducing falls, decoupling pervasive age-related systemic decline from overt functional impairment.

The impact of levodopa on the temporal components of gait in individuals with Parkinson’s disease (PD) is characterized by a significant restoration of rhythmic timing and a reduction in the bradykinetic elements of walking. Levodopa therapy primarily improves gait and pace-related gait measure. Specifically, the administration of levodopa has been shown to significantly increase cadence while simultaneously decreasing step duration, effectively allowing for a more fluid and rapid stepping frequency^15,19^. While these improvements are robust, it is noted that while levodopa optimizes the basic timing of steps, it may not fully resolve gait variability or postural instability to the same degree as it does for velocity and cadence^15^.

In conclusion, this study serves as a proof-of-concept, demonstrating that the dopaminergic system may act as a resilience mechanism for managing gait impairments in the elderly. Our findings show the improvement of cadence and step durations in older adults with MPS in L-dopa active group compared to placebo control group, supporting the hypothesis that levodopa treatment can improve gait in older adults with MPS.

## Limitations

A limiting factor of the study is the small sample size, which we attempted to overcome by employing a mixed linear model analysis, which allowed us to incorporate multiple pre-treatment outcome measure observations to better characterize the baseline gait characteristics within our sample. Furthermore, despite the randomization with a control group, persistent placebo effects cannot be ruled out. Replicating these findings with a larger sample size in a double-blind placebo controlled clinical phase 2a trial is necessary to verify the validity of these findings.

## Data Availability

All data produced in the present study are available upon reasonable request to the authors

