## Supplemental documents for "Effect of levodopa treatment on gait in older adults with mild parkinsonian signs"

**Supplementary Materials**

S1. Gait Factor Analysis Methods

To reduce the dimensionality of our gait dataset, factor analysis was applied to 14 gait variables across both participant groups at baseline (pre-treatment; N=14). A vector of all variable means and standard deviations was computed and used to center and scale the baseline dataset. The number of retained factors was identified using a scree plot and a minimal eigenvalue criterion of 1.5. A varimax rotation was applied to the loading score matrix. The rotated loading score matrix was thresholded so that all variable loadings with an absolute value less than 0.7 were set to 0, retaining only the strongest variable loadings.

Both baseline and follow-up (post-treatment; N=14) observations were centered and scaled using means and standard deviations previously obtained from baseline observations. Factor scores for both baseline and follow-up observations were obtained by taking a dot product of the centered and scaled data matrix with the rotated and thresholded loading score matrix.

S2. Gait Factor Analysis Results

A three-factor solution was selected for dimensionality reduction based on an eigenvalue cutoff of 1.5 (see supplementary figure 1). Based on the pattern and directionality of factor loading scores (see supplementary figure 2), the three gait factors were identified as “gait efficiency” (RC1), “gait rhythmicity” (RC2), and “gait turning” (RC3). Gait efficiency captures relative time spent during the more energetically efficient swing/single-support phase vs. stance/double-support phase of the gait cycle^20^. Gait rhythmicity captures how quickly steps are taken^21^. Gait turning captures the duration and the angle of the turning phase during the iSAW assessment.

**Supplementary Figure 1.** Factor analysis scree plot.
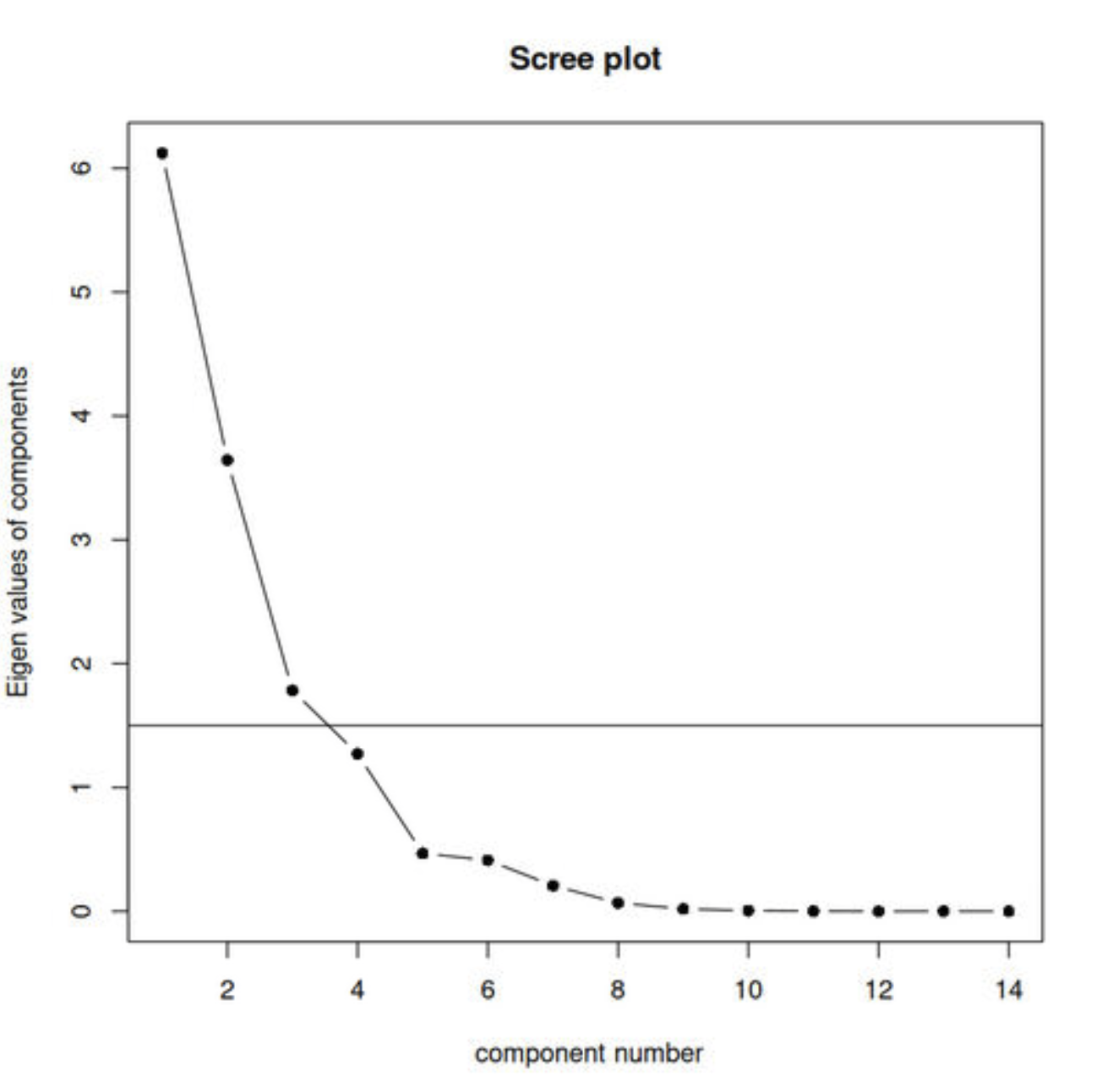

**Supplementary Figure 2.** Heatmap of factor analysis loading scores. Red colors reflect positive loading scores unto a gait factor (increase proportional to the factor score) and blue colors reflect negative loading scores unto a gait factor (decrease proportional to the factor score).

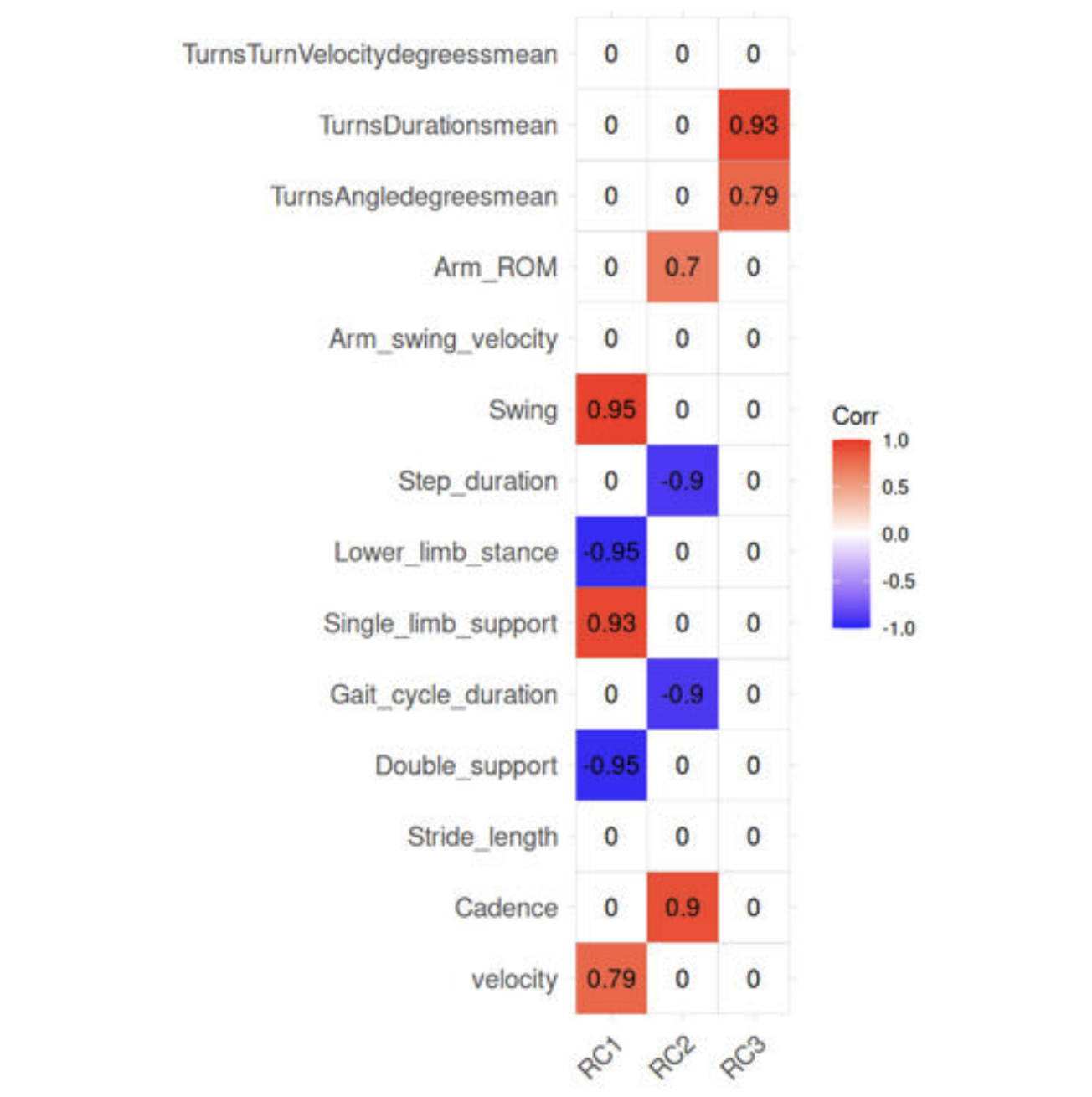
